# Approach to Machine Learning for Extraction of Real-World Data Variables from Electronic Health Records

**DOI:** 10.1101/2023.03.02.23286522

**Authors:** Blythe Adamson, Michael Waskom, Auriane Blarre, Jonathan Kelly, Konstantin Krismer, Sheila Nemeth, James Gippetti, John Ritten, Katherine Harrison, George Ho, Robin Linzmayer, Tarun Bansal, Samuel Wilkinson, Guy Amster, Evan Estola, Corey M. Benedum, Erin Fidyk, Melissa Estevez, Will Shapiro, Aaron B. Cohen

## Abstract

**Background:** As artificial intelligence (AI) continues to advance with breakthroughs in natural language processing (NLP) and machine learning (ML), such as the development of models like OpenAI’s ChatGPT, new opportunities are emerging for efficient curation of electronic health records (EHR) into real-world data (RWD) for evidence generation in oncology. Our objective is to describe the research and development of industry methods to promote transparency and explainability.

**Methods:** We applied NLP with ML techniques to train, validate, and test the extraction of information from unstructured documents (eg, clinician notes, radiology reports, lab reports, etc.) to output a set of structured variables required for RWD analysis. This research used a nationwide electronic health record (EHR)-derived database. Models were selected based on performance. Variables curated with an approach using ML extraction are those where the value is determined solely based on an ML model (ie, not confirmed by abstraction), which identifies key information from visit notes and documents. These models do not predict future events or infer missing information.

**Results:** We developed an approach using NLP and ML for extraction of clinically meaningful information from unstructured EHR documents and found high performance of output variables compared with variables curated by manually abstracted data. These extraction methods resulted in research-ready variables including initial cancer diagnosis with date, advanced/metastatic diagnosis with date, disease stage, histology, smoking status, surgery status with date, biomarker test results with dates, and oral treatments with dates.

**Conclusions:** NLP and ML enable the extraction of retrospective clinical data in EHR with speed and scalability to help researchers learn from the experience of every person with cancer.

## INTRODUCTION

A barrier to generating robust real-world evidence (RWE) is access to research-ready datasets that demonstrate sufficient recency, clinical depth, provenance, completeness, representativeness and usability. For studies using routinely collected electronic health record (EHR)-derived data, a considerable amount of data pre-processing and labor-intensive curation is required to create a dataset with clinically meaningful variables and outcomes needed for analysis (**Figure 1**).

**Figure 1.**
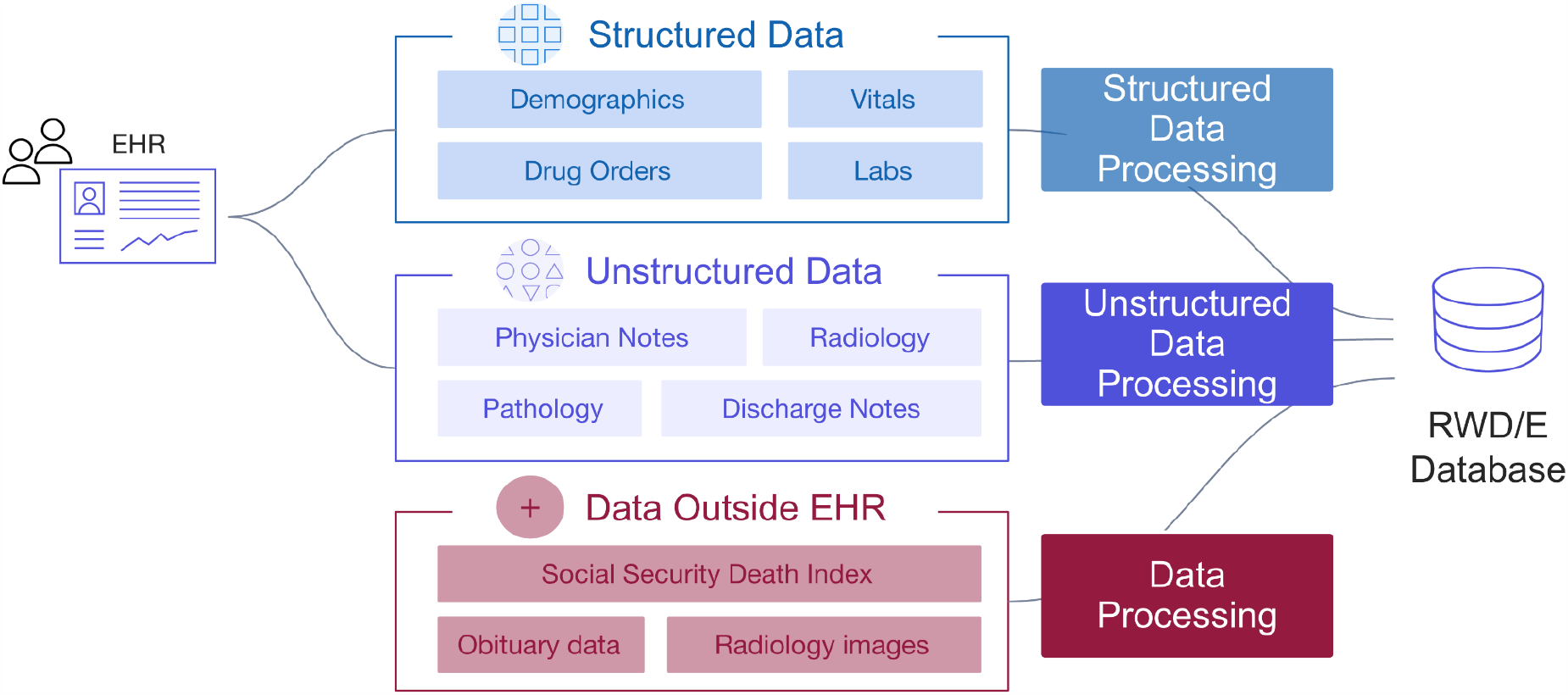
Overview of data variables defined by structured and unstructured information in HER.

The challenge is that so much valuable information is trapped within unstructured documents like clinician notes or scanned faxes of lab reports, where extracting the relevant data is far from trivial. The traditional approach to having clinical experts manually review patient charts to abstract data is time consuming and resource intensive.^1^ This approach limits the number of patients available for research purposes. Learnings can quickly become outdated—for example as new biomarkers and treatments emerge, the standards of care change, or new indicators for social determinants of health are prioritized. In other instances, answers to important research questions remain infeasible due to limited sample sizes.

Artificial intelligence (AI) advances in the areas of natural language processing (NLP) and machine learning (ML) have created new opportunities to improve the scale, flexibility, and efficiency of curating of high-quality real world data (RWD) in oncology.^2 3 4 5 6 7 8 9 10^ The definitions of foundational AI/ML terminology are provided in **Box 1**. When using ML and NLP for RWE, current guidance emphasizes transparency.^11 12 13 14 15^ The UK National Institute for Health and Care Excellence instructs that “where human abstraction or artificial intelligence tools are used to construct variables from unstructured data, the methods and processes used should be clearly described”.^11^

In response to guidance, the objective of this paper is to describe the general approach for applied NLP and ML methods that are used by Flatiron Health to extract data from unstructured documents stored in oncology care EHR. A key distinction in our terminology is the use of “abstraction” meaning performed by humans and “extraction” meaning performed by models.

Out of scope for this paper are other AI, ML, and NLP innovations and contributions from Flatiron Health, such as: model-assisted cohort selection^1 16^; continuous bias monitoring software^17^; automated mapping of laboratory data^18^; prediction of future health events^19^; and point-of-care products to improve patient care and clinical trials.^20 21^

## METHODS

### Overview

We developed a set of research analysis variables using information from the documents available in patient charts. Variables were selected when commonly required for retrospective observational studies in oncology but not consistently available in claims data or structured EHR data, where a corresponding version was curated by expert abstraction with a large amount of abstracted data available for training models.^22^

The variables curated through our ML extraction approach are those where the values are solely derived from the identification of clinical details in the EHR documents by an ML model in combination of NLP techniques and rules-based logic. It is important to note that these values are not predictions or inferences, but rather a direct extraction of information that is clearly documented in the EHR.

### EHR-derived Data Source

This study used the nationwide Flatiron Health EHR-derived de-identified database. The Flatiron Health database is a longitudinal database, comprising de-identified patient-level structured and unstructured data.^23 1^ At the time of this research, the database included de-identified data from approximately 280 US cancer practices (∼800 distinct sites of care).

Structured and unstructured data modalities are available in the database. EHR structured data elements include, but are not limited to, documented demographics (eg, year of birth, sex, race/ethnicity, etc.), vitals (eg, height, weight, temperature, etc.), visits, labs, practice information, diagnosis codes, medication orders, medication administrations, ECOG performance status, health insurance coverage, and telemedicine (**Figure 1**). EHR unstructured data and documents include, but are not limited to, paragraphs of clinic visit notes, PDF scans of lab results, radiology images with reports, pathology reports, and communications between the patient and care team (**Figure 2**). For the purpose of this paper, all the figures contain fictional representations of documents, sentences, dates and patient IDs.

**Figure 2.**
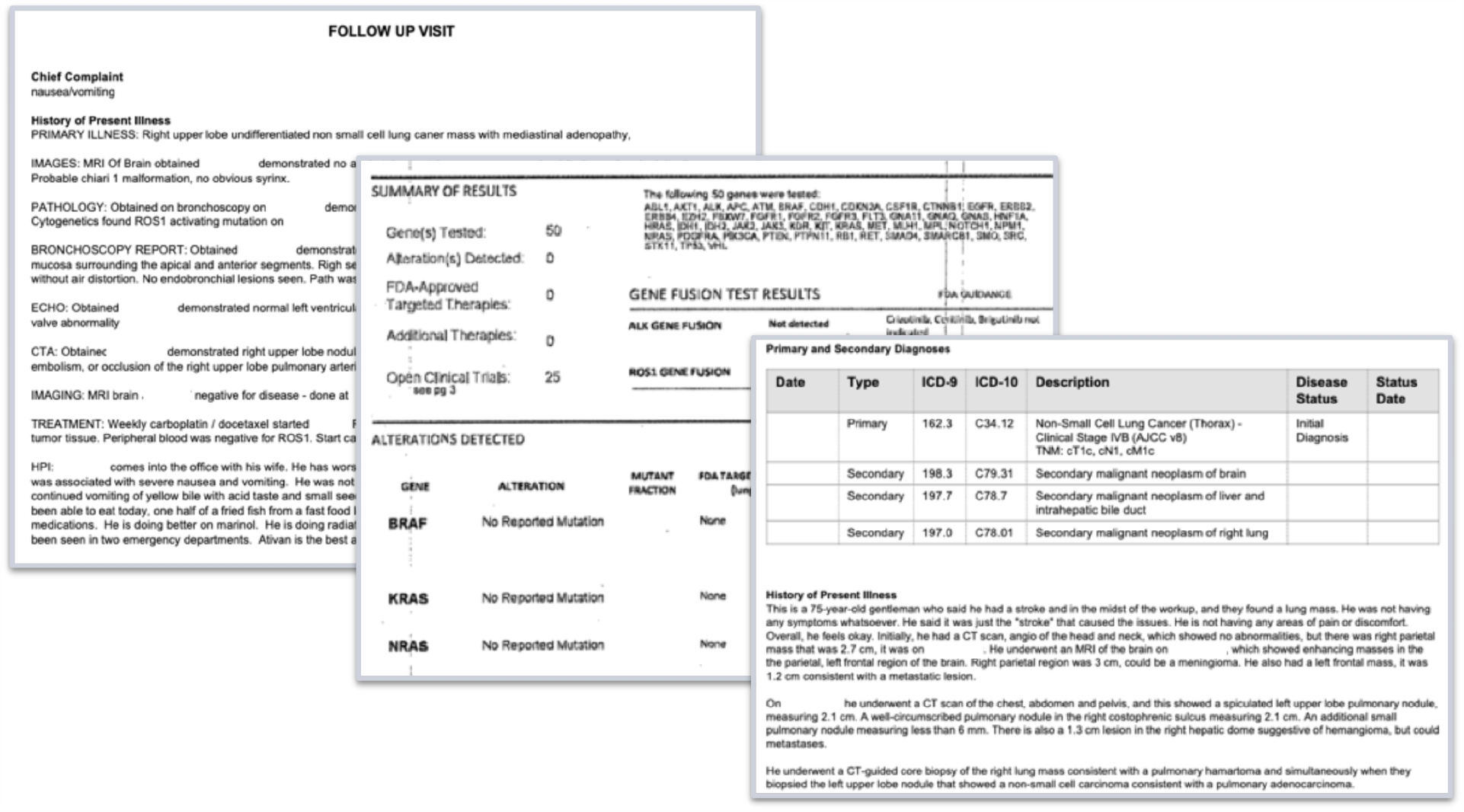
Examples of unstructured documents from EHR that are used as inputs for ML-extraction of information (all dates and patient IDs are fictitious).

### Patient Population

The large general cross-tumor cohort includes all patients with at least one International Classification of Diseases (ICD)-9 or ICD-10 cancer code and at least one unique-date clinic encounter documented in the EHR (reflected by records of vital signs, treatment administration, and/or laboratory tests) on or after January 1, 2011. The distribution of patients across community and academic practices largely reflects patterns of care in the US, where most patients are treated in community clinics, but can vary between cancer types.

### Clinical Expert Abstraction of Variables for Model Development

Critical information in patient charts has been manually abstracted by trained clinical experts (ie, clinical oncology nurses or tumor registrars), following a set of standardized policies and procedures. To abstract data from patient charts, we use a foundational technology^24^ that enables clinical experts to more easily review hundreds of pages of documents to determine patient characteristics, treatments, and outcomes documented in the EHR (**Figure 3**).

**Figure 3.**
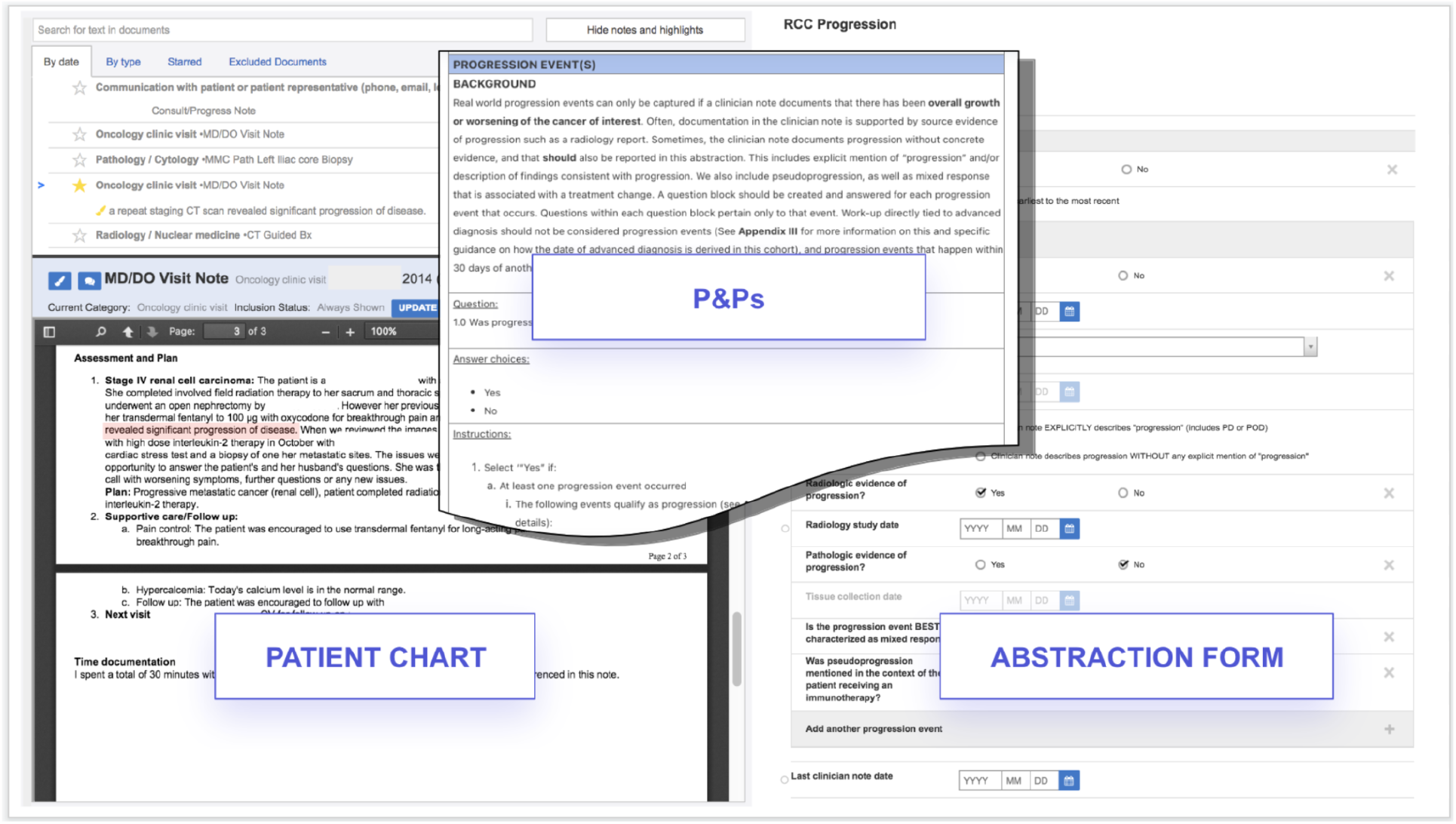
Technology enabled expert abstraction. Abbreviations: P&Ps, Policies and Procedures. All dates and patient IDs are fictitious.

Years of manual abstraction by a workforce of thousands of abstractors at Flatiron Health have created a large and high-quality corpus of labeled oncology EHR data. Clinically-relevant details specific to each cancer type are abstracted from every form of clinical documentation available in the EHR, including clinic visit notes, radiology reports, pathology reports, etc. Abstractors are trained to locate and document relevant information by following policies and procedures tested and optimized for reliability and reproducibility through iterative processes, and oversight is provided by medical oncologists.

The abstraction process undergoes continuous auditing to monitor abstractor performance, while proprietary technology links each curated data point to its source documentation within the EHR, enabling subsequent review. At the individual patient level, this approach provides a recent and robust longitudinal view into the clinical course, capturing new clinical information as it is documented within the EHR.

Flatiron Health has abstracted sets of clinically meaningful variables from more than 300,000 people with cancer to develop disease-specific de-identified research-ready databases.^23^ Limited by the capacity of human abstractors, there had remained millions of patients with cancer in the Flatiron Health database for whom no unstructured data had yet been curated to create variables with the clinical depth needed to generate meaningful insights. If a hypothetical variable required 30 minutes of chart review by a clinical expert to abstract the information of interest for 1 patient, then it would take a team of 100 full-time abstractors more than 7 years to finish defining 1 variable for a population of 3 million patients.

### Overview of Machine Learning Extraction Approach

The objective of this application of NLP and ML methods was to replicate the expert abstraction process described in the previous section. When developing ML models for extracting information, all of the clinical abstractor expertise that was incorporated into the manual abstraction of variables is available to learn from through training. Once iterated upon and placed in production, ML models can automate information extraction from unstructured clinical data sources in a way that mimics expert clinical abstractors. Flatiron Health has developed a collection of proprietary AI/ML algorithms that include, but are not limited to, deep learning architectures,^25^ text snippet-based modeling approaches,^26^ and extraction of patient events and dates. ^27 28 29^

Alongside the manually-abstracted labels, we use NLP to pull relevant textual information from charts to use as inputs to train built-for-purpose ML models and model architectures for a given extraction task. Through this process we can make our end variables appropriate for disease-specific or pan-tumor (ie, histology-independent) applications. For example, by deciding whether or not to use model training data sourced from curated disease-specific cohorts or any-cancer cohorts, we can make our model’s output variables built-for-purpose to be used in an analysis that generates meaningful RWE for a specific research question.

A range of model architectures were evaluated and considered for the purpose of information extraction for variables of interest. The model output of variable classes ranged, including:

- binary (eg, metastatic diagnosis Yes/No)
- categorical unordered (eg, never smoker, history of smoking, current smoker)
- categorical ordered (eg, cancer stage I-IV)
- date (eg, 02/05/2019 start of oral treatment X)

Date and classification can come from the same model, separate models, or connected models.

### Natural Language Processing to Generate Model Inputs

For each variable of interest, we begin with clinical experts constructing a list of clinical terms and phrases related to the variable. Since models are trying to extract explicit information from charts, rather than infer it, only terms that are directly relevant to a specific variable are included (eg, when extracting a patient’s histology, terms could include “histology,” “squamous,” and/or “adenocarcinoma,” but do not include treatment or testing terms from which the histology might be indirectly inferred).

Next, we use NLP techniques to identify sentences in relevant unstructured EHR documents (eg, oncology visit notes, lab reports, etc.) that contain a match to one of the clinical terms or phrases. The contextual information surrounding the clinical term is critical because the words at the beginning of a sentence may change the interpretation of a key word at the end of a sentence. ML models can understand if the clinical concept appears and under what context—such as, if negativity, speculation, or affirmation exists in the surrounding clinical terms (ie, snippets). Where applicable, any associated dates within these sentences are also identified. These sentences are then transformed into a mathematical representation that the model can interpret. The output of this document processing is a broad set of features aimed at fully capturing document structure, chronology, and clinical terms or phrases.

### Machine Learning Model Development

#### Features and Labels

The features defined by NLP become the inputs provided to the model to score the likelihood that a given patient document is associated with each class of a particular categorical variable (eg, histology categories of non–squamous cell carcinoma, squamous cell carcinoma, non–small cell lung cancer [NSCLC] histology not otherwise specified). The final model output is the variable value for each patient. The labeled dataset is commonly split into three subsets: a training set, a validation set, and a test set. The training and validation sets are used to build the model, which often involves an iterative development process, while the test set is used to evaluate the performance of the final ML model.

#### Model Development

The training set comprises labeled data points that are used to optimize the model’s parameter values. In an iterative process, training examples are provided to the model, its outputs are compared to the labels, and the parameter values are adjusted in response to errors. By using manually-abstracted values as labels, the objective of this process is for the model to learn what answer a human abstractor would give when reading a specific clinical text.

The validation set is used to assess how well the model has learned these associations. Because the model does not see any data from patients in the validation examples during training, they can be used to estimate how it will perform on new, unlabeled examples once it is put into production. Validation performance is commonly assessed using metrics such as precision, recall, and F1 score (See Box 1 Key Terms in Machine Learning). These aggregate metrics, combined with review of individual errors, inform decisions about search terms, text preprocessing steps, and model architectures. Experimentation continues until a final “best” model is identified.

When a ML model is trained to perform a classification task, it outputs scores for each possible class for each data point. These scores are between 0 and 1 and show the probability that a patient belongs to each class, based on information in their electronic health record. However, the scores may vary if the wording in the records is unusual or if there is conflicting information. For example, if a patient’s cancer stage is being restaged, there may be multiple mentions of different stages in the record, and the model may assign moderate scores to each stage if the restaging event is unclear.

To produce a discrete class value, the class with the highest score is often chosen, but other approaches may optimize performance. In particular, a probability threshold may be chosen such that a patient will be classified into one class if and only if their score exceeds the threshold. The optimal threshold depends on factors such as class balance and is typically chosen empirically.^30^ When no class receives a sufficiently high score, another option is to defer to abstraction to resolve uncertainty (Waskom et al, in press, 2023).

We explored and experimented with a range of ML models and architectures for the purpose of extracting specific variable information from the EHR. Deep learning architectures included long short-term memory (LSTM), Gated recurrent units (GRU), and bidirectional encoder representations from transformers (BERT).^31 32 33^ These models can learn thousands or millions of parameters, which enable them to capture subtleties in the text. They read sentences as a whole and use the words around a clinical term to incorporate surrounding context when determining the extracted class. When they receive very large texts as inputs, they can figure out where the relevant information is and focus on this section and its context.

For example, in LSTMs, words are passed into the model sequentially; during each step through a sentence, the model has access to memory (ie, an internal state) that is impacted by the previous word, in effect allowing the model to “remember” the previous word (Figure 4). The LSTM block combines the new word with the information that came before to derive a more contextually rich representation of the word. For instance, when the LSTM reads the word “Advanced,” it remembers (via the model’s internal state) that it was preceded by the word “not” and is more likely to classify the patient as “not advanced.”

**Figure 4.**
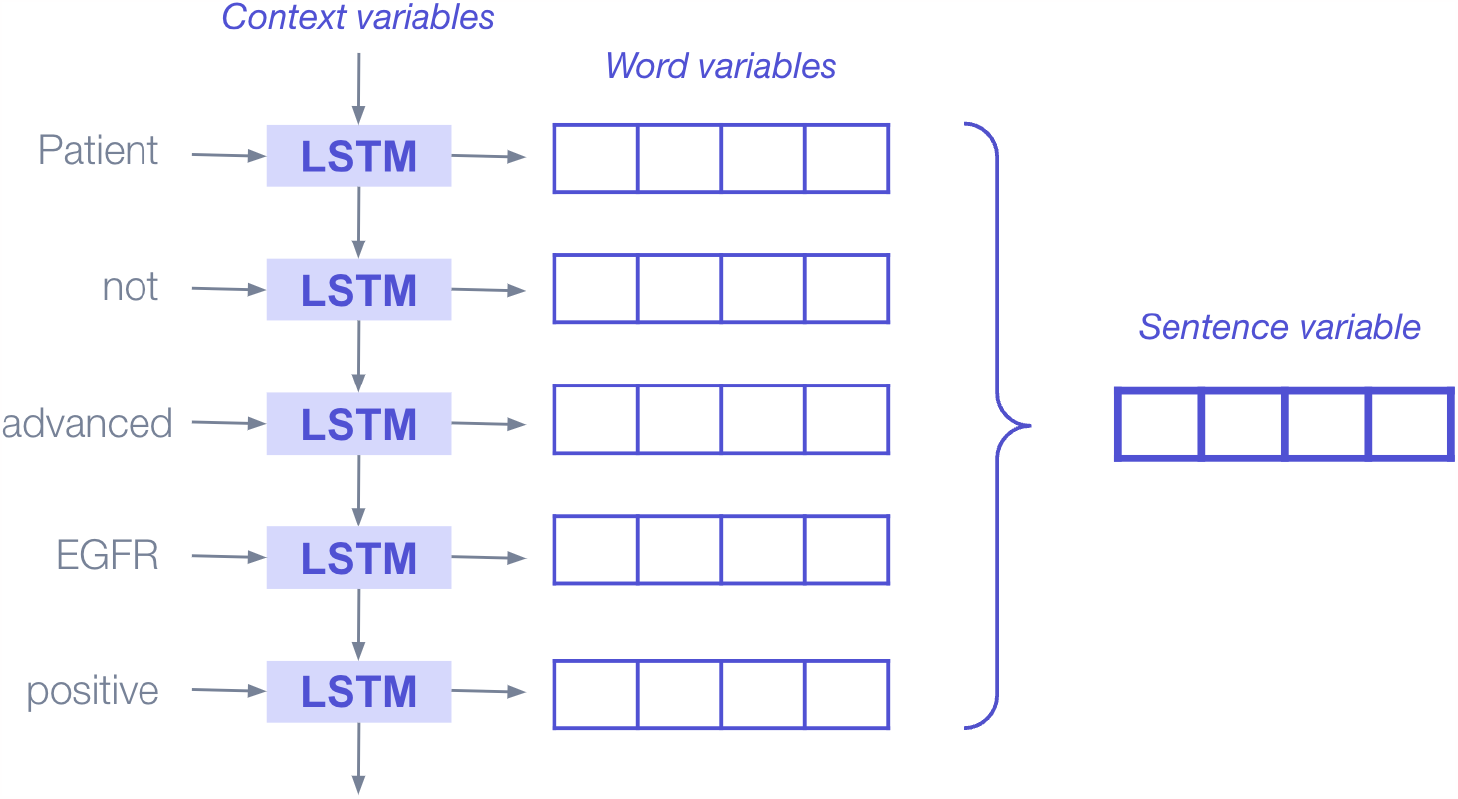
Illustration of deep learning bidirectional LSTM blocks applied sequentially to produce representations (aka, embeddings or encodings) that encapsulate the information added to the sentence by each new word. Abbreviations: LSTM, long short-term memory.

### Model Evaluation and Performance Assessment

Once iteration on the ML model is complete, final model performance is measured on a test set that uses manually-abstracted labels as the source of truth. Test sets are designed to be large enough to power both top-level metrics and sub-group stratifications on a “held out” set, that is, on data not used to train the ML model or validate performance during prototyping. This allows the test set to assess the model’s ability to correctly classify data points that it has never seen before, which is typically referred to as the “generalization” of the model.

Measuring performance is a complex challenge because even a model with good overall performance might systematically underperform on a particular subcohort of interest, and because while conventional metrics apply to individual models, dozens of ML extracted variables may be combined to answer a specific research question. We use a research-centric evaluation framework^34^ to assess the quality of variables curated with ML. Evaluations include one or more of the following strategies: (1) overall performance assessment, (2) stratified performance assessment, and (3) quantitative error analysis, and (4) replication analysis. As variables curated with NLP and ML are expected to be incorporated into the evidence generated that will guide downstream decision-making, variable evaluation can also include replication of analyses originally performed using abstracted data. Replication analyses allow us to determine whether ML-extracted data—either individual variables or entire datasets—are fit-for-purpose in specific use cases by assessing whether they would lead to similar conclusions.

Specific variable-level performance metrics are only interpretable for cohorts with characteristics that are similar to the test set, depending on inclusion criteria such as the type and stage of cancer. As a result, we do not report them here.

Python was the primary coding language used in the development of ML models described here. Institutional Review Board approval of the study protocol was obtained before study conduct, and included a waiver of informed consent.

## RESULTS

We successfully extracted key information from unstructured documents in the EHR using the developed proprietary ML models trained on large quantities of data labeled by expert abstractors. For this paper, we are focusing the results on examples within NSCLC as they were the first applications we developed. A set of 10 ML models output 20 distinct RWD variables for analysis, including initial cancer diagnosis with date, advanced/metastatic diagnosis with date, disease stage, histology, smoking status, surgery details, biomarker test results, and oral treatments with dates. Language snippets were the inputs for these models to produce a data point for each patient for each variable as outputs, illustrated in **Figure 5**.

**Figure 5.**
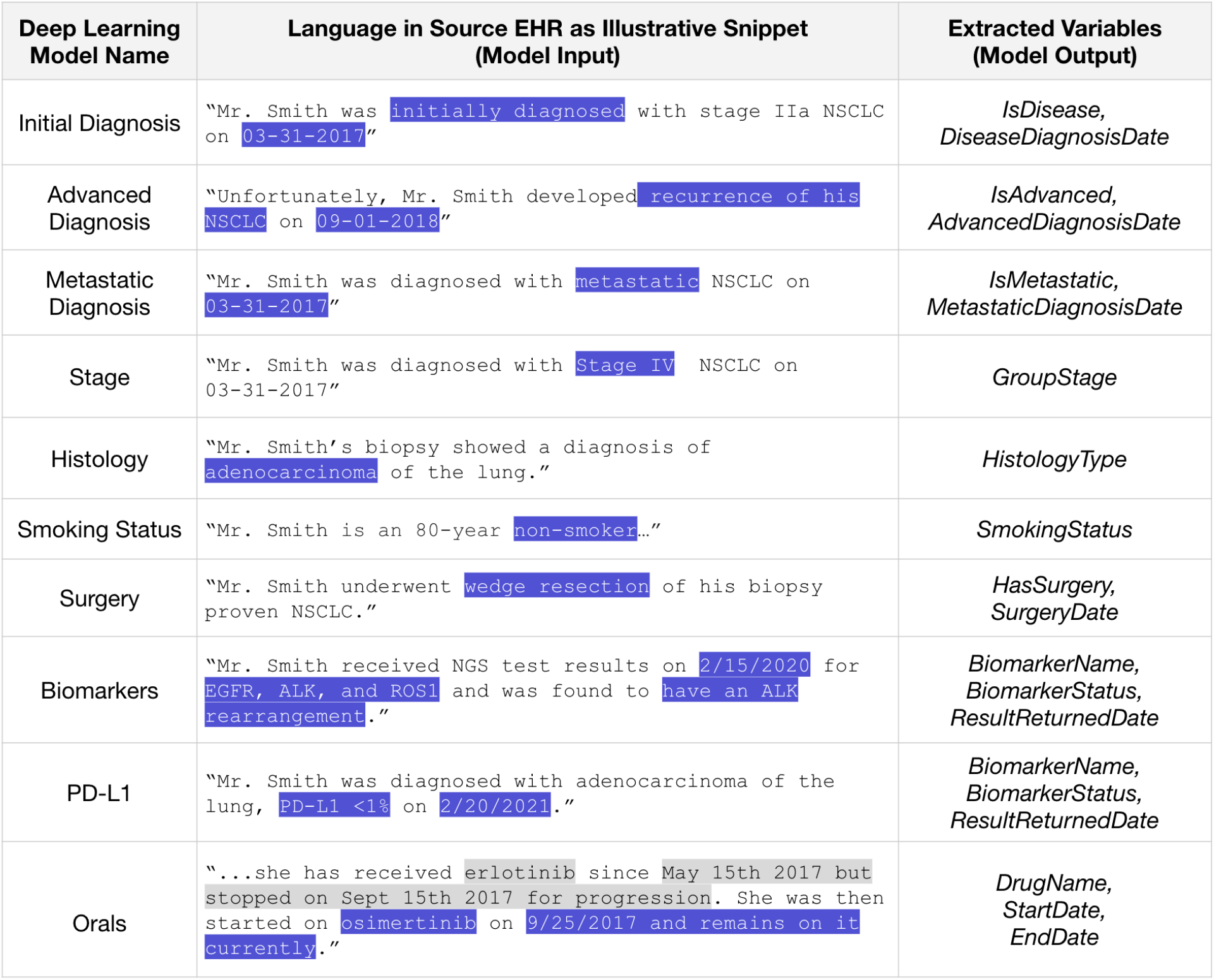
Sentences (fictional examples here) from EHR are inputs to deep learning models that produce a data variable value for each patient as an output. Language snippets are only extracted around key terms from which a variable might be extracted, and not around terms from which it could be indirectly inferred. Abbreviations: EHR, electronic health record; PD-L1, programmed death ligand 1. All dates and patient IDs are fictitious.

Datatables containing variables curated by an approach using ML had the same appearance and functionality as variables curated with an approach using technology-enabled expert human abstraction (**Figure 6**).

**Figure 6.**
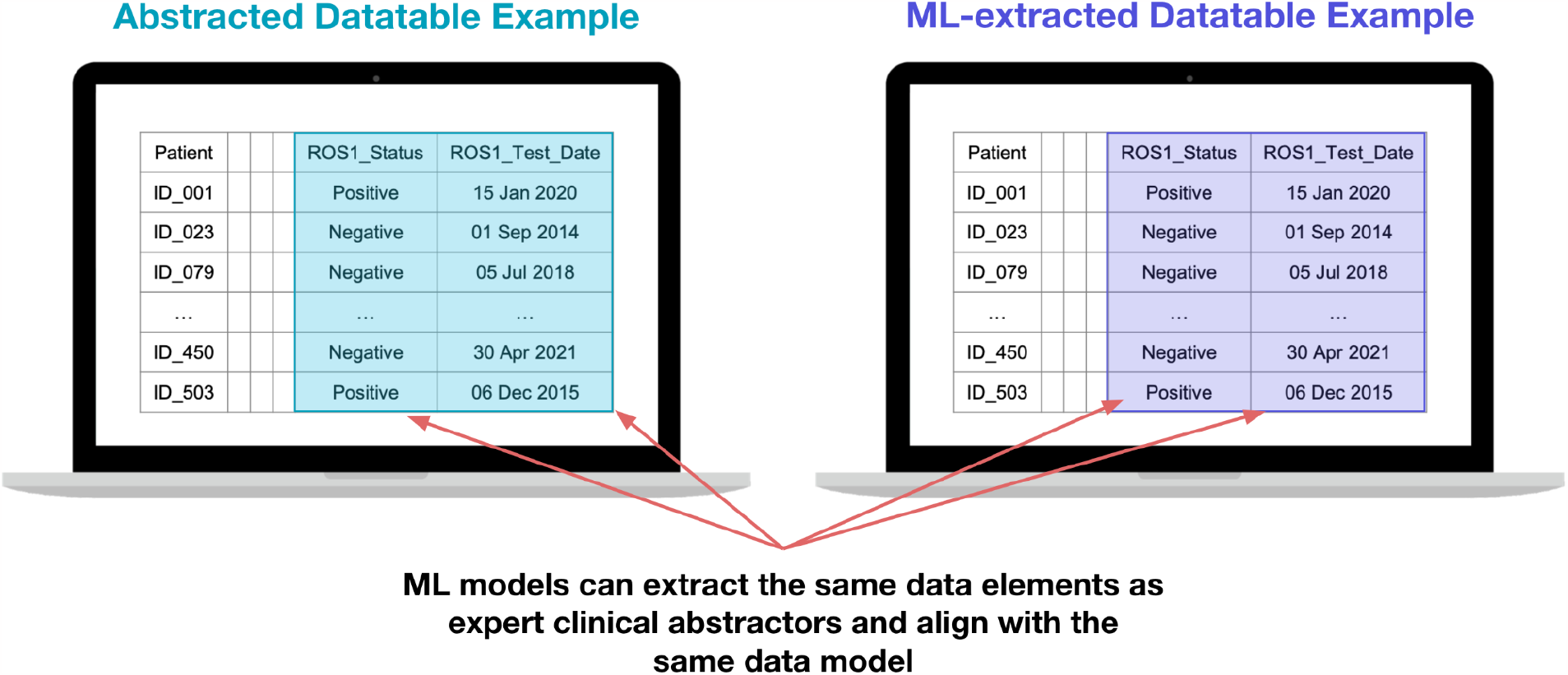
Illustration of a data table with variables curated by an approach using expert human abstractors (right) alongside a data table with variables curated by an approach using deep learning models (left) shows opportunity for exchangeable utility in real-world data analysis. All dates and patient IDs are fictitious.

Models had high performance when trained for disease-specific applications and histology-independent (ie, tumor agnostic) patient cohorts. Detailed performance metrics are out of scope for this paper. Beyond satisfactory ML metrics, we found that in some cases ML-extraction can achieve similar error rates as manual abstraction by clinical experts (Waskom et al, in press, 2023), and replication studies suggest that research analysis relying on multiple variables can reach similar results and conclusions when using variables curated by ML-extraction compared with human experts (Benedum et al, in press, 2023).^35 36^

Approaches and learnings related to specific variables are described below.

### Application Examples

We have developed ML models for a number of different variables and use cases. A few of the more prominent models and their associated use cases are described below.

#### Cancer Diagnosis and Dates

We successfully developed deep learning models focused on the task of extracting initial, advanced, and metastatic cancer diagnosis and the corresponding diagnosis dates. Historically, ICD codes have been used as a proxy for diagnosis, as they are well captured in structured EHR data due to their use in billing. However, we have seen that the precision of ICD codes varies by disease, is not strongly correlated with disease prevalence in the larger population, and can be lower than 50%. With that in mind, extracting accurate diagnosis information is imperative to understanding patient populations, as errors at the diagnosis level propagate to all other variables. These models build on prior foundational research on extracting information from longitudinal clinic notes.^37 38^ A conceptual diagram of the approach is presented in **Figure 7**. The initial, advanced, and metastatic variables are generated using multiple, distinct ML models. We have found success chaining the models together—providing the output of one model as the input to the next—to prevent conflicting predictions and improve overall accuracy. An early investigation into model performance has been presented previously.^39^

**Figure 7.**
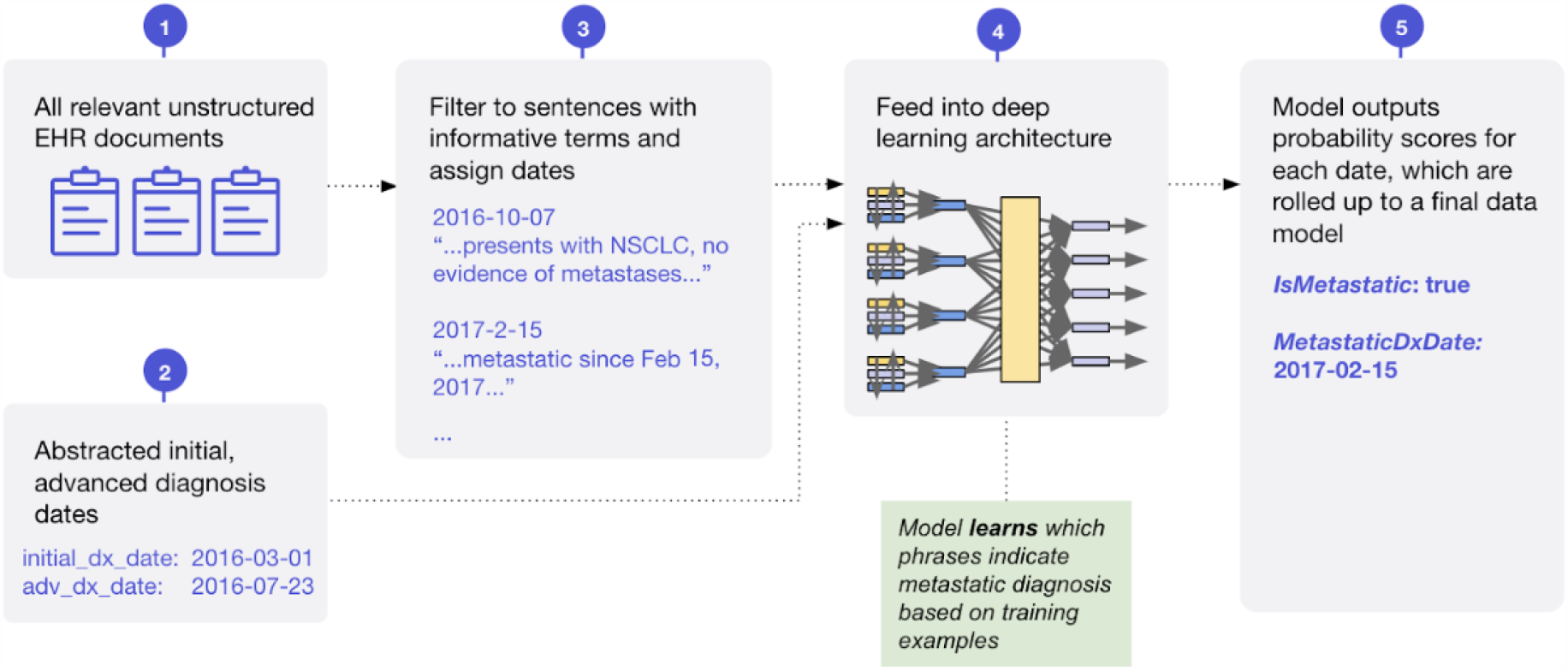
Conceptual diagram of machine learning model for extraction of metastatic diagnosis and date.

Additional complexity exists when trying to identify patients with rare cancers, primarily due to the low number of labels. We have developed techniques to allow our models to successfully generalize to these diseases, with few or no labels provided during training from the target disease(s).

#### Disease Stage and Histology

We successfully developed a deep learning model to extract cancer stage information and a second ML model to extract the histology of the tumor. One example of how we used this approach for a disease-specific application was training on patients with NSCLC. This model was designed to extract main stage (I-IV) and substage (letters A-C) granularity. Histology was extracted as a non-ordered categorical variable with the possible variable values of non–squamous cell carcinoma, squamous cell carcinoma, or NSCLC histology not otherwise specified.

As cancer stage is documented similarly across solid tumor diseases, we were able to scale our approach to extract disease stage in a tumor-agnostic cohort with a similar deep learning architecture but training data composed of patients with multiple cancer types. While hematologic cancers have some important differences from solid organ cancers when it comes to assigning stage (risk stratification scores, no concept of metastatic disease, etc.), we found success using a deep learning model to extract this information for a number of hematologic cancers. Tumor histology is not as straightforward to scale across cancer types, as different cancers originate from different possible cell types (and therefore have different histologies). This means that to date, we use distinct histology models for each type of cancer. Performance evaluations for disease stage and histology are conducted at each category level and by cancer type as appropriate for use cases.

#### Smoking Status

We successfully developed a deep learning model to extract information in the patient chart that indicates whether or not the patient has any lifetime history of smoking. The categorical variable output has the possible values as history of smoking, no history of smoking, or unknown. The most relevant sentences for this model were most often found in social history paragraphs of text that is a standard section in clinical encounter notes. Critical document categories that enabled high accuracy of this model included access to oncology clinic visit notes, radiology reports, surgery reports, lab reports, and pulmonary test result reports. The smoking status model was trained on a broad dataset of patients that included many cancer types for whom we have abstracted smoking status.

#### Surgery and Surgery Date

We successfully developed a deep learning model to extract information about whether the patient had a primary surgical procedure where the intent was to resect the primary tumor. As these types of surgeries often happen in outpatient facilities or hospitals, this valuable documentation lives in unstructured text formats in the oncology EHR. We have abstracted surgery data in certain disease cohorts but, because of the similarity in documentation approaches across cancer types, we were able to train a model that is tumor agnostic. This allowed us to scale surgery status and date in larger patient populations and in new disease types.

#### Biomarker Testing Results and Result Date

We successfully developed and deployed models to generate variables for biomarker testing, including extraction of the dates that the patient had results returned (**Figure 6**). One part of the model is able to identify whether or not a given document for a patient contains a biomarker test result. A separate part of the model is able to extract from the document the date a result was returned and the biomarker result. Early efforts with a regularized logistic regression model were presented previously^40^ and more sophisticated models have been developed since.

A model first cycles through every EHR document for a given patient to understand whether or not the document contains biomarker testing results. These models rely on access to lab reports, including those saved in the EHR as a PDF or image of a scanned fax. The models are able to process report documents produced by different labs (eg, Foundation Medicine, Caris, Tempus, etc.) in addition to the clinician interpretations in visit notes.

A separate model then extracts the biomarker (eg, included but not limited to *ALK, BRAF, EGFR, KRAS, MET, NTRK, RET, ROS1*, or PD-L1) and test result (eg, positive, negative, or unknown). This approach gives our ML models flexibility to extract biomarkers that the model may not have seen before in training. For PD-L1, where results are quantitatively reported, a separate ML model was developed to extract percent staining, with classes of <1%, 1%-49%, ≥49%, and unknown.

Since patients can receive biomarker testing multiple times throughout the treatment journey and at multiple facilities, it is possible that a given patient has more than one biomarker test result and date. For each patient, this allows us to determine biomarker status at different clinical milestones (eg, advanced diagnosis date, start of second-line treatment, etc.).

#### Oral Treatments and Treatment Dates

We successfully developed a deep learning model to extract oral treatment information, including the treatment name, and the span for which the treatment was administered. In contrast to intravenous therapies such as chemotherapy or immunotherapy in which each dose is ordered and administered to be given in the clinic or infusion room, oral therapies are prescribed to patients to be filled by an outpatient pharmacy, which is frequently outside the clinic site. To have a complete understanding of all cancer treatments received or delayed (eg, postponed during a hospitalization), it is necessary to enumerate the use of oral treatments through review of unstructured clinician visit notes, prescriptions, and communications with the patient or patient representative. Important information to select within the paragraphs of text include the treatment name, start date, and end date. We previously published an initial framework^38^ for extracting drug intervals from longitudinal clinic notes, prescriptions, and patient communication documents and have developed more sophisticated and accurate methods since then. We found the visit notes contained key pieces of information about treatments being held or started when patients were hospitalized.

The model is trained to select mentions of a specific list of drug names used for oral treatment in the specific cancer type, along with the start date and end date. These oral treatment variables are generated using three distinct ML models. The list of oral treatments of interest were specific to each disease and defined by oncology clinicians. Expert abstraction from charts includes collection of treatment start dates and discontinuation dates that were used for ML model training.

## DISCUSSION

This paper described one approach to curating real-world oncology data variables from unstructured information in EHR using NLP and ML methods. Model development was possible with access to a large and high quality corpus of labeled oncology EHR data produced via manual abstraction by a workforce of thousands of clinical expert abstractors over the course of several years. We now have models that are able to meet or even exceed human abstraction performance on certain tasks. Using a performance evaluation framework^34^ for variables curated using the approach of ML extraction we affirmed high quality and fitness-for-use in RWE generation. We have shown that validations using the combination of multiple ML-extracted variables in one RWD analysis demonstrated no meaningful difference in RWE findings based on replications with the Flatiron Health variables curated by ML extraction compared with expert human abstraction (Benedum et al, in press, 2023).^36 35^

Crucial information about clinical details may be recorded only within free-text notes or summaries in unstructured EHR documents. Our models primarily rely on deep learning architectures, because curating data from such sources usually requires techniques that capture the nuances of natural language. At the same time, we select model architectures on a case-by-case basis depending on what works best for each variable, and we have found that the quality of the training data and labels can be just as if not more important to success. Progress in AI research is rapidly accelerating, as demonstrated by the impressive generative abilities of models like gpt3 and its ChatGPT application. We expect that future advances will likewise make deeper and more nuanced clinical concepts accessible to ML extraction, although the generative framework itself may remain more suited for tasks such as summarization^41^ than for scalable curation of structured real-world datasets.

The mission to improve and extend lives by learning from the experience of every person with cancer is more important than ever. With increasingly specific combinations of patient characteristics, disease, and therapy, we need to learn from as many relevant examples as possible to have statistically meaningful results. ML expands the opportunity to learn from patients who have been oppressed or historically marginalized in oncology clinical trials.^42 43^ As oncology care rapidly evolves, and the treatment landscape becomes more personalized—targeting new biomarkers, finely tuned to increasingly particular patient profiles—transparent fit-for-purpose applications of ML will have increasing importance. With high performance models, we can truly learn from every patient, not just a sample. It also creates an opportunity to improve the completeness of RWD variables that were previously defined by only structured data elements, reducing potential bias in evidence.

There are strengths and limitations to the EHR curation approaches described here. Strengths include the large size, representativeness, and quality of training data used; success across a multitude of cancer types; and the explainability of approach to finding clinical details in documents. Massive volumes of high-quality expert abstracted data were a unique advantage for training high-quality ML models. Researchers at Stanford have confirmed similar capabilities with a different EHR dataset—detecting the timeline of metastatic recurrence of breast cancer.^44^ The ML models described here were trained for and applied only in a US population.^23^ While the most suitable model architectures for each variable may be transferable across country borders, a limitation of this approach is that models must be re-trained with local data for highest performance.

The capability to build ML models that can extract RWD variables accurately for a large number of patients further enables the possible breadth and depth of timely evidence generation to answer key policy questions and understand the effects of new treatment on health outcomes.

## Data Availability

The data that support the findings of this study have been originated by Flatiron Health, Inc. Requests for data sharing by license or by permission for the specific purpose of replicating results in this manuscript can be submitted to.

## Acknowledgements

The authors would like to thank Selen Bozkurt, Sharang Phadke, Shreyas Lakhtakia, Nick Altieri, Qianyu Yuan, Geetu Ambwani, Lauren Dyson, Chengsheng Jiang, Somnath Sarkar, Javier Jimenez, Arjun Sondhi, Alexander Rich, Benjamin Birnbaum, Andrej Rosic, Barry Leybovich, Jamie Irvine, Nisha Singh, and Sankeerth Garapati. Catherine Au-Yeung and Tanya Elshahawi contributed to illustration design.

## Author contributions

BA, JK, SN, and EE contributed to the conception of this review paper. AB, GH, GA, JK, JG, JR, KH, KK, MW, RL, TB, SW developed the ML models. CB, ME, EF, AC, and BA conducted performance evaluations and validations. BA wrote the first draft of the manuscript. SN, GA, WS, MW, ME, AB, AC, EF, RL wrote sections of the manuscript. All authors contributed to manuscript revision, read, and approved the submitted version.

## Abbreviations

AI: artificial intelligence
BERT: bidirectional encoder representations from transformers
EHR: electronic health records
LSTM: long term short memory
ML: machine learning
NPV: negative predictive value
NSCLC: non–small cell lung cancer
P&Ps: Policies and Procedures
PPV: positive predictive value
RWD: real world data
RWE: real-world evidence

## Box 1.

Key Terms in Machine Learning

### Foundational machine learning (ML) definitions

- **Class:** One of the possible values that a binary or categorical variable can take.
- **Labels:** The known classes associated with data used to train or evaluate an ML model.
- **ML-Extracted:** Algorithmic extraction of data from documented evidence in the patient chart (either structured or unstructured) at the time of running the model. Techniques include ML and NLP, in contrast to other data processing methods such as abstraction or derivation.
- **Model:** An ML algorithm with a specific architecture and learned parameters that takes inputs (eg, text) and produces outputs (eg, extracted diagnosis).
- **Natural Language Processing (NLP):** A field of computational systems (including but not limited to ML algorithms) that enable computers to analyze, understand, derive meaning from, and make use of human language
- **Score:** A continuous output from a model that can be interpreted as the model-assigned probability that a data point belongs to a specific class.
- **Threshold:** A cutoff value that defines classes when applied to continuous scores. Binary variables (eg, whether a patient has had surgery) have a natural default threshold of 0.5, but different thresholds might be leveraged depending on the relative tolerance for false positives vs. false negatives required.

### Performance metric definitions

- **Sensitivity (Recall):** The proportion of patients abstracted as having a value of a variable (ie, group stage = IV) that are also ML-extracted as having the same value.
- **PPV (Precision):** The proportion of patients ML-extracted as having a value of a variable (ie, group stage = IV) that are also human abstracted as having the same value.
- **Specificity:** The proportion of patients abstracted as not having a value of a variable (ie, group stage does not = IV) that are also ML-extracted as not having the same value.
- **NPV:** The proportion of patients ML-extracted as not having a value of a variable (ie, group stage does not = IV) that are also abstracted as not having the same value.
- **Accuracy:** The proportion of patients where the ML-extracted and abstracted values are identical. For variables with more than 2 unique values (eg, group stage), accuracy within each class is calculated by binarizing the predictions (eg, for Accuracy of group_stage = IV, all abstracted and ML-extracted values would be defined as either “IV” or “not IV”.
- **F1 Score:** Computed as the harmonic mean of sensitivity and PPV. For a binary classifier, the threshold that maximizes F1 can be considered the optimal balance of sensitivity and PPV.

